# A Scoping Review Protocol on Integration of mobile-linked POC diagnostics in community-based healthcare: User Experience

**DOI:** 10.1101/2022.10.14.22281115

**Authors:** Siphesihle R. Nxele, Boitumelo Moetlhoa, Kabelo Kgarosi, Tivani Mashamba-Thompson

## Abstract

**Background:** Mobile-linked point-of-care diagnostics forms an integral part of diagnostic health services for efficient communication between patients and healthcare professionals despite geographical location and time of diagnosis. The efficiency of this technology lies in the user experience which means that the interaction of the user with the implemented technology needs to be simple, convenient, and consistent. Having a well-structured user experience of these devices in community-based healthcare will aid in sustainable implementation. Herein, we propose to conduct a literature search to systematically map out evidence based on mobile-linked POC diagnostics user experience at a community level in resource-limited settings.

**Methodology:** The proposed scoping review will be guided by the advanced Arksey and O’Malley methodological framework and further advanced by Levac *et al*. A comprehensive search will be conducted to find relevant published literature from the following electronic databases: Scopus, Web of Science, EBSCOhost (Medline, CINAHL, Africa-wide, Academic Search Complete). Grey literature will also be searched, including reports from government and international organizations such as World Health Organization (WHO), Foundation for Innovative New Diagnostics (FIND), and the Food and Drug Administration (FDA). Two independent reviewers will screen the relevant studies and the degree of the agreement will be determined by calculating Cohen’s kappa statistic. The quality of eligible data will also be appraised using the mixed method appraisal tool version 2018.

**Discussion:** We anticipate that the planned scoping review will present useful evidence to inform stakeholders on the integration of mobile-linked diagnostic devices in community-based healthcare which will guide further research on the subject.

## 1. Background

Mobile health (mHealth) refers to the use of mobile devices with medical-based software to make healthcare more accessible to patients and potentially improve the manner in which patient medical information is managed (1). mHealth is important in the delivery of diagnostic health services in order to bridge the accessibility gap by allowing two-way communication between patients and healthcare professionals despite geographical location and time (2). The integration of mobile-linked point-of-care (POC) diagnostic tools in community-based healthcare has made medical care accessible to patients and clinical staff in a cost-effective and efficient manner (3), shortening the time between diagnosis and treatment of patients. Resource-limited communities, however, still face issues such as poor infrastructure and lack of resources for advanced medical care, thus motivating the need for enhanced POC diagnostic tools such as mHealth.

In order to address this shortfall, the Joint United Nations Programme on HIV/AIDS (UNAIDs) and World Health Organization (WHO) developed a strategic plan that entailed the linking of wireless mobile communication to POC diagnostics and the implementation of this strategy in health policies and programs, which was aimed at addressing the advanced technology improvements and their contribution to healthcare systems (4). The estimated number of smartphone users in resource-limited settings such as Sub-Saharan Africa was nearly 36% in 2018, and this is predicted to rise to about 66% by 2025, making it a powerful (5). The WHO further compiled a mHealth assessment and planning scale (MAPS) toolkit (6) for the advancement of discussions surrounding the upscaling of mHealth innovations for women, children, and adolescent health which emphasized the need for further research and development within the field of mHelath and its accessibility in under-resourced communities. Successful implementation and integration of such technologies require a deeper understanding of technological experiences, lifestyle, and general behaviours of the user (7).

In community-based healthcare, the integration of mobile-linked POC diagnostics needs to be carried out in a manner that will be useful and accessible in resource-limited settings (8). Reports have shown that the implementation of mHealth should make patients more knowledgeable about their health condition and can create a more efficient way for health care professionals with different expertise to liaise and make more informed medical decisions and diagnoses (9). In both regards, the aim should be for this technology to be user-friendly, efficient on the patient and medical expert end, and secure so doctor-patient confidentiality is maintained. It is also important to understand user experience regarding how the implementation of mobile-linked POC diagnostic tools will affect the workflow of medical personnel (10). User experience in the context of mHealth technologies looks at focusing the design of the technology to the target user so that the product can be used effectively and efficiently, while achieving user satisfaction in a specific context (11). Evidence of user experience in the currently used mobile-linked POC diagnostics at a community level in resource-limited settings is unclear. The proposed scoping review aims to systematically map evidence on mobile-linked POC diagnostics user experience at a community level in resource-limited settings. It is anticipated that the findings of this proposed scoping review will help guide future research on improving the usability of mHealth-linked POC diagnostics and improving access to diagnostics services in settings that have limited access to laboratory services.

## 2. Methodology

The methodological framework that was proposed by Arksey and O’Malley (12) and further advanced by Levac et al., (13) will guide this scoping review in five stages. These guiding stages will be as follows: (i) identify the research question; (ii) identify relevant studies; (iii) select eligible studies; (iv) selection of sources of evidence and (v) ethical considerations. Preferred Reporting Items for Systematic reviews and Meta-Analyses for scoping reviews (PRISMA-ScR) guidelines will be used to present the review results (14). In compliance with the National (South Africa) Open Science Policy (15), this protocol is registered on Open Science Framework and accessible via this link: https://archive.org/details/osf-registrations-zs3pb-v1

### i. Identification of a research question

We employed the PCC (Population, Concept, and Context) nomenclature to determine the eligibility of the research question for a scoping review (**Table 1**). The research question is: What is the level of user experience of mobile-linked (m-linked) POC diagnostics in community-based healthcare? The identified population was m-linked POC diagnostics which we define as technology that allows for the screening and diagnostics of communicable and non-communicable diseases in remote settings by healthcare professionals and patients (4) such as Covid-19 tests (16) and chest x-ray evaluations (17). This technology would aid in shortening the time between testing and clinical diagnosis (18). The concept of this study has been identified as user experience in health technologies which refers to taking into consideration the target community’s values, desires, expectations, personal objectives, and lived experience when implementing such technologies (19) and the context identified as community-based healthcare. Community-based healthcare refers to improving healthcare in targeted populations, which involves providing healthcare services on a local, personalized level, where without this strategy, communities would not have access to appropriate medical care (20). An example of community-based healthcare would include community healthcare facilities that provide services such as community nursing, aged care, and occupational therapist services (21).

**Table 1:**
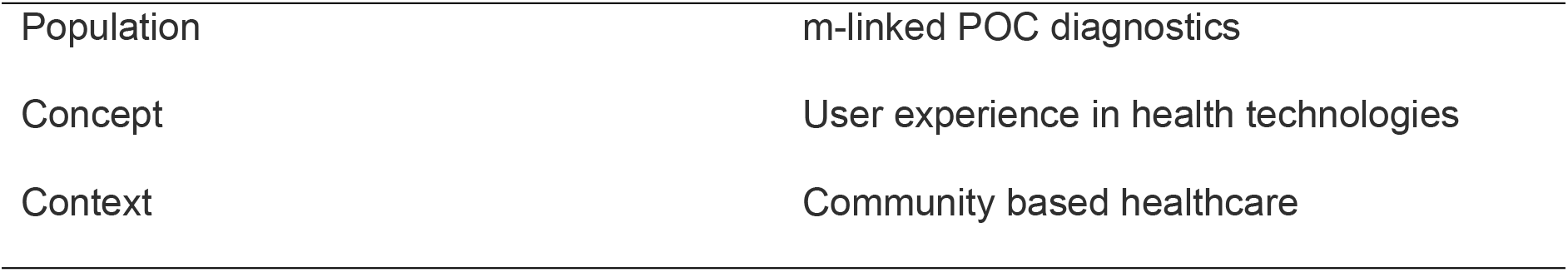
PCC for determining the eligibility of the research question

### (ii) Identification of relevant studies

The following databases will be used to conduct a comprehensive and reproducible literature search using the following electronic databases: Scopus, Web of Science, EBSCOhost (Medline, CINAHL, Africa-Wide, Academic Search Complete). The principal investigator (PI), subject specialist, and university librarian will develop a comprehensive search strategy to ensure the correct use of indexing terminology and Medical Subject Headings (MeSH) terms. Grey literature including dissertations/theses, conference proceedings, websites of international organizations such as WHO, and government reports, will also be explored. Additional relevant studies will be identified by manually searching all references cited in the included studies to identify studies not indexed in electronic databases. There will be no language restrictions applied, to minimize the risk of excluding relevant studies. The following keywords will be used (and refined to suit each database): 1- “User experience” or “user experience in health technologies” or “UX” or “UX in health technologies” 2- “point of care” or “point of care testing” or “point of care diagnostics” or “point of care diagnostic services” or “mobile-linked point-of-care diagnostics” and 3- “community-based health” or “community-based healthcare” or “primary healthcare”. Each search will be documented in detail, showing the keywords date of search, electronic database, and the number of retrieved studies, and the results of the search will be tabulated. The search strategy will be optimized by adopting the search summary table (SST) outlined by Bethel et al., (22) as a guide. The SST will be used to improve and report on the effectiveness of the search strategy.

A pilot search in various electronic databases was conducted to demonstrate the feasibility of addressing our research question using a scoping review. The results of our pilot search are indicated in **Table 2**.

**Table 2:** Results of pilot search in selected electronic databases

| Date of search | Database | Keywords | Search results |
| --- | --- | --- | --- |
| 29/09/2022 | SCOPUS | ( TITLE-ABS-KEY ( "mobile-linked point-of-care testing" OR "mobile linked point-of-care diagnostics" OR "digital-linked point-of-care diagnostics" OR "point-of-care diagnostics" OR "Diagnostic Test" OR "point-of-care testing" OR "Point-of-Care Systems" OR "Diagnostic Test" OR point-of-care OR "user experience" OR "UX" OR "usability" OR "personalized medicine" OR "community based healthcare" OR "community health" ) ) | 259 |
| 5/10/2022 | MEDLINE | ((MM "Mobile Diagnostic Test Kits Approval") OR (MM "Home Diagnostic Tests") OR (MM " mobile-linked point-of-care testing ") OR " mobile point-of-care diagnostics" OR "digital-linked point-of-care diagnostics" OR "point-of-care testing" OR "Diagnostic Test" OR "point-of-care systems" OR "Point-of-Care Systems" OR "Diagnostic Test" OR Point-Of-Care OR POCT) OR User Experience OR UX OR Community based healthcare OR community health) | 223 |
| 5/10/2022 | CINAHL | ((MM "Mobile Diagnostic Test Kits Approval") OR (MM "Home Diagnostic Tests") OR (MM " mobile-linked point-of-care testing ") OR " user experience of mobile point-of-care diagnostics" OR "point-of-care testing" OR "Diagnostic Test" OR "point-of-care systems" OR "Point-of-Care Systems" OR "Diagnostic Test" OR Point-Of-Care OR POCT) OR User Experience OR UX OR Community based healthcare OR community health) | 106 |
| 5/10/2022 | Africa Wide | "User experience in mobile-linked point-of-care diagnostics" | 22 |
| 6/10/2022 | Academic search complete | ( TITLE-ABS-KEY ( "mobile point-of-care testing" OR "point-of-care testing" OR "Diagnostic Test" OR "point-of-care systems" OR "Point-of-Care Systems" OR "digital-linked point-of-care diagnostics" OR "Diagnostic Test user experience" OR user experience OR UX ) AND TITLE-ABS-KEY ( "real time connectivity ease of specimen collection environmental friendliness affordable sensitive specific user friendly rapid equipment free delivered" OR reassured OR "community based healthcare diagnostics" OR "community health" OR "user experience" ) ) | 173 |
| 6/10/2022 | Web of Science | "mobile point-of-care testing" OR "Diagnostic Test" OR "point-of-care systems user experience" OR "real time connectivity ease of specimen collection environmental friendliness affordable sensitive specific user friendly rapid equipment free delivered" OR user experience OR "UX" OR "community-based healthcare" OR "community health diagnostics" | 198 |
| **TOTAL** |  |  | **981** |

### (iii) Selection of eligible studies

This scoping review will be guided using inclusion and exclusion criteria to ensure that relevant studies are selected:

#### a. Inclusion criteria

The included articles should meet the following criteria:

- Articles reporting on mobile-linked POC diagnostics in a community-based healthcare setting
- Articles reporting on technology that allows for the screening and diagnostics of communicable and non-communicable diseases in remote settings
- Articles reporting on the user experience (UX) in health technologies
- Articles reporting on the evidence of community-based healthcare on a local, personalized level
- All reviews providing evidence of mobile-linked point-of-care diagnostic systems

#### b. Exclusion criteria

Articles were excluded from the scoping review if they fall under the following description:

- Articles that lacked evidence on point-of-care diagnostics in community-based healthcare
- Articles that lacked evidence on the user experience (UX) of health technologies at the primary healthcare level

All reviews not providing evidence of integrated mobile-linked point-of-care diagnostic systems

### (iv) Selection of sources of evidence

The articles will be screened in three stages: title, abstract and full article. Reviewers will use a screening tool developed by the PI and pilot tested by the reviewers. The eligible articles will be exported to an Endnote 20 library, and the duplicates will be removed. The PI will screen abstracts in parallel with the co-reviewer. After screening the abstracts, the reviewers will discuss any discrepancies in the selected articles until a consensus is reached. Two reviewers will then screen the full texts of articles selected during the first stage. A third screener will resolve any discrepancies in the selected articles after the full-text screening. Both abstract and full-article screening will be guided by the screening tool that factored in all aspects of the inclusion/exclusion criteria and the PCC elements. The level of agreement between screeners’ results after screening the abstracts and full articles will be determined by calculating Cohen’s kappa statistics. The kappa statistics were interpreted as follows: values <0.1 indicate no agreement and 0.10–0.20 indicate none to slight, 0.21–0.40 as fair, 0.41–0.60 as moderate, 0.61–0.80 as substantial, and 0.81–1.00 as almost perfect agreement. Database search and screening results will be presented using the PRISMA chart (Figure 1).

#### a. Charting the data

A data charting form will be used to capture all relevant data from the articles reviewed. Two independent reviewers will pilot the data charting form and recommend modifications that will be implemented. The following data will be extracted from the included articles: author and year of publication, the title of study, aim of the study, study design, study setting, study population, type of point-of-care test investigated, the level of user experience of the identified POC diagnostic tests and other significant findings.

#### b. Collating, summarizing, and reporting the results

The quantitative data extracted from the included articles will be presented in a table and graphic format. Qualitative data will be summarised according to emerging themes.

#### c. Quality appraisal

The quality of the included articles will be evaluated using the mixed method appraisal tool (23) version 2018 (24). Using MMAT, the methodological quality of five categories of research will be appraised, which include the following: qualitative research, randomized controlled trials, nonrandomized studies, quantitative descriptive studies, and mixed methods studies.

The quality appraisal process will be carried out by two independent reviewers. The following percentage scores will be used to grade the quality of evidence where: (i) ≤50% represents low-quality evidence, (ii) 51–75% represents average-quality evidence, and (iii) 76–100% represents high-quality evidence.

### (v) Ethical considerations

This scoping review will be developed and compiled using existing literature. No primary data will be collected to answer the research question.

## Discussion

According to the Foundation for Innovative New Diagnostics (FIND) 2021 strategy report, no diagnostic tests exist for 60% of the pathogens already identified by the World Health Organization (WHO) to have the potential of developing into another pandemic In addition, half of the known top 20 diseases which are responsible for the highest mortality rate have no reliable diagnostic test kits (25). To solve the setbacks, the successful and sustainable implementation of reliable mobile linked POC diagnostic tools will depend on the user experience in terms of accessibility, acceptability, and adoption by the target users (7). With no consideration of user experience, the implementation of mobile linked POC diagnostics will be short-lived. The user experience (UX) will play a big role to achieve sustainable implementation and use.

In order to gather relevant evidence to answer our research question, this scoping review will exclude articles that lack evidence on point-of-care diagnostics in community-based healthcare. Articles that lack evidence on the user experience (UX) of health technologies in community-based healthcare and all reviews that do not provide evidence of integrated mobile-linked point-of-care diagnostic systems will also be excluded. The third sustainable development goal (SDG3) is to ensure healthy lives and promote well-being for all. More specifically, SDG 3.4 aims to reduce premature mortality by a third, from non-communicable diseases through prevention and treatment (26). This work will be published in peer-reviewed journals and presented to relevant stakeholders to achieve the above-outlined goals on a global scale.

## Data Availability

No datasets were generated or analysed during the current study. All relevant data from this study will be made available upon study completion.

## Acknowledgments

The authors would like to extend their appreciation to the University of Pretoria Faculty of Health Sciences Library services for their assistance with optimizing the search strategy.

## Bibliography

1. Källander K, Tibenderana JK, Akpogheneta OJ, Strachan DL, Hill Z, ten Asbroek AH, et al. Mobile health (mHealth) approaches and lessons for increased performance and retention of community health workers in low-and middle-income countries: a review. Journal of medical Internet research. 2013;15(1):e2130.

2. Hurst EJ. Evolutions in telemedicine: from smoke signals to mobile health solutions. Journal of Hospital Librarianship. 2016;16(2):174–85.

3. Mechael PN. The case for mHealth in developing countries. Innovations: Technology, Governance, Globalization. 2009;4(1):103–18.

4. Osei E, Kuupiel D, Vezi PN, Mashamba-Thompson TP. Mapping evidence of mobile health technologies for disease diagnosis and treatment support by health workers in sub-Saharan Africa: a scoping review. BMC Medical Informatics and Decision Making. 2021;21(1):1–18.

5. GSMA. The Mobile Economy. Sub-Saharan Africa 2020. GMSA. 2020.

6. Organization WH. The MAPS toolkit: mHealth assessment and planning for scale: World Health Organization; 2015.

7. Portz JD, Bayliss EA, Bull S, Boxer RS, Bekelman DB, Gleason K, et al. Using the technology acceptance model to explore user experience, intent to use, and use behavior of a patient portal among older adults with multiple chronic conditions: descriptive qualitative study. Journal of medical Internet research. 2019;21(4):e11604.

8. Heidt B, Siqueira WF, Eersels K, Diliën H, van Grinsven B, Fujiwara RT, et al. Point of care diagnostics in resource-limited settings: A review of the present and future of PoC in its most needed environment. Biosensors. 2020;10(10):133.

9. Kirkscey R. mHealth apps for older adults: a method for development and user experience design evaluation. Journal of Technical Writing and Communication. 2021;51(2):199–217.

10. Oinas-Kukkonen H, Räisänen T, Leiviskä K, Seppänen M, Kallio M. Understanding the role of user experience for mobile healthcare. New technologies for advancing healthcare and clinical practices: IGI Global; 2011. p. 169–81.

11. Harte R, Glynn L, Rodríguez-Molinero A, Baker PM, Scharf T, Quinlan LR, et al. A human-centered design methodology to enhance the usability, human factors, and user experience of connected health systems: a three-phase methodology. JMIR human factors. 2017;4(1):e5443.

12. Arksey H, O’Malley L. Scoping studies: towards a methodological framework. International journal of social research methodology. 2005;8(1):19–32.

13. Levac D, Colquhoun H, O’Brien KK. Scoping studies: advancing the methodology. Implementation science. 2010;5(1):1–9.

14. Tricco AC, Lillie E, Zarin W, O’Brien K, Colquhoun H, Kastner M, et al. A scoping review on the conduct and reporting of scoping reviews. BMC medical research methodology. 2016;16(1):1–10.

15. Bezuidenhout L, Gould C, Farrant J. Academy of Science of South Africa launches a mapping survey of life science research and diagnostic activity in South Africa. SAMJ: South African Medical Journal. 2013;103(7):0-.

16. Zhang HM, Dimitrov D, Simpson L, Singh B, Plaks N, Penny S, et al. A web-based, mobile responsive application to screen healthcare workers for COVID symptoms: Descriptive study. MedRxiv. 2020.

17. Schwartz AB, Siddiqui G, Barbieri JS, Akhtar AL, Kim W, Littman-Quinn R, et al. The accuracy of mobile teleradiology in the evaluation of chest X-rays. Journal of telemedicine and telecare. 2014;20(8):460–3.

18. Nayak S, Blumenfeld NR, Laksanasopin T, Sia SK. Point-of-care diagnostics: recent developments in a connected age. Analytical chemistry. 2017;89(1):102–23.

19. Pravettoni G, Triberti S. P5 eHealth: An agenda for the health technologies of the future: Springer Nature; 2020.

20. Maraccini AM, Galiatsatos P, Harper M, Slonim AD. Creating clarity: Distinguishing between community and population health. American Journal of Accountable Care. 2017;5(3):27–32.

21. Wiggers J, McElwaine K, Freund M, Campbell L, Bowman J, Wye P, et al. Increasing the provision of preventive care by community healthcare services: a stepped wedge implementation trial. Implementation Science. 2017;12(1):1–14.

22. Bethel AC, Rogers M, Abbott R. Use of a search summary table to improve systematic review search methods, results, and efficiency. Journal of the Medical Library Association: JMLA. 2021;109(1):97.

23. Lo H, Frauendorf V, Wischke S, Schimmath-Deutrich C, Kersten M, Nuernberg M, et al. Ambulatory Use of Handheld Point-of-Care Ultrasound (HH-POCUS) in Rural Brandenburg - A Pilot Study. Ultraschall in der Medizin (Stuttgart, Germany : 1980). 2021.

24. Hong QN, Fàbregues S, Bartlett G, Boardman F, Cargo M, Dagenais P, et al. The Mixed Methods Appraisal Tool (MMAT) version 2018 for information professionals and researchers. Education for information. 2018;34(4):285–91.

25. Rodriguez B, Brock W, Saran K. THE ESSENTIAL ROLE OF DIAGNOSTICS IN GLOBAL HEALTH & DEVELOPMENT. 2022.

26. Organization WH. Report of the first meeting of the Strategic and Technical Advisory Group for Noncommunicable Diseases: virtual meeting, 27–28 October 2021. 2021.

